# The Forgotten Tract of Vision in Multiple Sclerosis: Vertical Occipital Fasciculus, Its Integrity, and Visuospatial Memory

**DOI:** 10.1101/2021.01.25.21250430

**Authors:** AmirHussein Abdoalizadeh, Soheil Mohammadi, Mohammad Hadi Aarabi

## Abstract

**Background:** Visual disturbances are a common disease manifestation and a major patient complaint in Multiple Sclerosis (MS) due to lesions damaging white matter tracts involved in vision. Vertical Occipital Fasciculus (VOF) connects ventral and dorsal visual streams and was neglected for more than a century. It has recently become under focus in brain-related disorders. Thus, its role in the visual dysfunction in MS needs to be clarified.

**Objective:** Evaluate the integrity of bilateral VOFs in MS and its association with clinical and visual evaluations.

**Methods:** 56 relapsing-remitting MS (RRMS) and 25 healthy controls (HC) were recruited. We acquired MS Functional Composite, Expanded Disability Status Scale (EDSS), and Brief Visuospatial Memory Test – Revised (BVMT-R), and structural and diffusion MRI scans. After VOF tractography, its integrity markers were statistically tested for between-group differences and clinical and visual tests associations.

**Results:** RRMS patients had lower fiber integrity in bilateral VOFs compared to HC. Lower integrity of bilateral VOFs was associated with poor clinical outcomes, higher visual score in EDSS, and lower total immediate and delayed recall in BVMT-R.

**Conclusion:** VOF damage is seen in RRMS and is associated with visual symptoms and visuospatial learning impairments.

## 1. Introduction

Multiple sclerosis (MS) is a chronic neurodegenerative disorder characterized by inflammatory demyelination of the central nervous system (CNS), generally presenting with periods of neurological deficits followed by episodes of remissions, called relapsing-remitting multiple sclerosis (RRMS)^1^. Visual symptoms are one of the initial manifestations of disease in nearly 25% of cases^2^, and have been attributed to lesions damaging the optic nerve causing optic neuritis^2^, or white matter tracts such as optic radiation^3^.

Diffusion-Weighted Imaging (DWI) can be used to derive tract-specific properties, such as fractional anisotropy (FA) and mean, radial, and axial diffusivity (MD, RD, AD, respectively). These metrics collectively provide information regarding the health, inflammation, or even myelination of neural fibers^4^. For example, a DWI study of the optic nerve in MS patients with optic neuritis shows elevated MD and reduced FA, indicating axonal loss through demyelination^5^. Moreover, DWI studies of optic radiation show a significant increase in MD, RD, and AD, accompanied by a decline in FA, indicating myelin loss and Wallerian degeneration^3, 6^.

Vertical occipital fasciculus (VOF) is regarded as the primary communicator between dorsal and ventral visual streams^7^. It was first described by Carl Wernicke in 1881 but was neglected for over a century until Yeatman et al. identified it using DWI tractography^8^. According to the substantial theory of ventral and dorsal visual streams, the ventral stream is involved in the interpretation of color^9^ and shape^10^, while the dorsal stream is involved in the interpretation of spatial comprehension^11^, motion, and disparity^7, 12^; thus, their connection via VOF to integrate both types of information seems inevitable.

Although the optic nerve, and the optic radiation have both been studied in MS, the role of VOF in the pathophysiology of visual disturbances in MS is obscure. In this study, we aim to shed light on the role of VOF in disabilities of MS by studying its integrity derived from DWI and its associations with clinical disability and visual disturbances.

## 2. Methods and Materials

### 2.1. Participants

57 RRMS patients and 26 healthy controls (HC) were recruited via Cross-Modal Research Initiative for Multiple Sclerosis and Optic Neuritis (CRIMSON), conducted in Multiple Sclerosis Research Center (MSRC), Sina Hospital, Tehran, Iran. In brief, all the patients undergo a battery of neuropsychiatric evaluations, blood sampling, and MRI. More information regarding the inclusion and exclusion criteria are explained in details elsewhere^13^. The CRIMSON study has been reviewed and accepted by the Ethical Review Board of Tehran University of Medical Sciences.

### 2.2. Evaluations

#### 2.2.1. Clinical Markers of Disease Severity

Expert clinicians evaluated the clinical status of patients based on the Expanded Disability Status Scale (EDSS)^14^ and MS Functional Composite (MSFC)^15^. EDSS involves clinically-oriented measures of disability evaluated by a clinician or self-report. MSFC is based on three tests itself: 9-hole peg test (9HP), timed 25-foot walk (25FW), and Paced Auditory Serial Addition Test (PASAT), to inclusively and objectively address several motor and cognitive aspects of patients.

#### 2.2.2. Visual

EDSS visual (optic) functions subscale (EDSS-Vis) was also used for optic function evaluation. It is scored based on clinical examination (Snellen chart and optical disc pallor) and patient self-report (presence of scotoma)^14^. Moreover, we used Brief Visuospatial Memory Test – Revised (BVMT-R) to assess visuospatial learning in our RRMS subjects^16^. In this test, six images are presented in a sheet for 10 seconds. Then the subject is asked to draw those images from memory. This trial is repeated three times, and after 25 minutes of doing other tasks the subject is asked to draw the images from memory. Each trial is scored if the location and shape of the drawn object (one point each) are same as the sheet presented (12 points/trial). Then, total immediate recall (BVMT-T), delayed recall (BVMT-DR), retained percentage (BVMT-R%), discriminability (BVMT-Dis), and response bias (BVMT-B) are calculated (Table 1). There were no BVMT evaluations available for HC in the CRIMSON study; however, a study by Eshaghi et al. in the same research center on another set of subjects has provided mean and standard deviation for BVMT-T and BVMT-DR for HCs^17^.

**Table 1.** Measures derived from Brief Visuospatial Memory Test – Revised, their procedure and scoring. BVMT = Brief Visuospatial Memory Test, Max = Maximum score of.

| Measure | Abbreviation | Procedure | Scoring |
| --- | --- | --- | --- |
| Delayed Recall | BVMT-DR | Draw the six images shown in trials 1 to 3 from memory after 25 minutes of doing other tasks. | Accurately drawn (one point) and correctly located (one point) for each image (total = 12 points). |
| Learning | BVMT-L |  | Max (Trials 2 or 3) – Score of trial 1 |
| Total Immediate Recall | BVMT-T |  | Sum of scores for trials 1 to 3 |
| Retained Percentage | BVMT-R% |  | BVMT-DR / Max (Trials 2 or 3) |
| Target Discriminability | BVMT-Dis | Twelve figures are shown to the subject to identify if the presented figure has been presented previously or not. | [True Positives – False Positives] |
| Response Bias | BVMT-B | Based on answers to BVMT-Dis procedure. | $(\text{No. of false positives} + 0.5) / (1 - [\text{corrected hit rate} - \text{corrected false alarm}])$ |

### 2.3. Imaging Acquisition

All the scans were acquired using a Siemens Avanto 1.5T scanner: A magnetization prepared rapid-acquisition gradient-echo (MPRAGE) T1 sequence [repetition time (TR) = 2730ms, echo time (TE) = 2.81ms, Inversion Time (TI) = 1000ms, field of view (FoV) = 256mm, voxel-size = 1×1×1 mm], a T2-weighted turbo spin echo with variable flip angle (TSE-VFL) [TR = 3200ms, TE = 473ms, FoV = 256mm, voxel-size = 1×1×1 mm], a Fluid-Attenuated Inversion Recovery (FLAIR) sequence [TR = 9400ms, TE = 83ms, TI = 2500ms, FoV = 250mm, voxel-size = 1.3×1.0×3.0 mm], and a DWI sequence [TR = 9500ms, TE = 93ms, voxel-size = 2×2×2.1 mm, b-value = 1000 s/mm^2^, in 64 diffusion directions] with 3 b0 sequences.

### 2.4. Imaging Analysis

#### 2.4.1. Lesion Filling

Lesions were manually filled on the FLAIR and T2 scans. Then, the T2-to-T1 registration warp was applied to bring lesions into the T1-space. The lesions were filled by the intensity of their nearby normal-appearing white matter^13^.

#### 2.4.2. DWI Analysis

For our imaging analysis, we used the cloud computing platform brainlife.io^18^. We used the mrtrix3 preprocess app comprising denoising, eddy-current, Gibbs ringing, and bias field correction on the unprocessed DWI data and lesion-filled T1 scans^19^. We then used the automated atlas-based tractography^20^ based on the HCP842 tractography atlas^21^ within DSI Studio (dsi-studio-atk.app). The automated tractography is based on the Hausdorff distance between each track based on atlas proposed by Yeh et al.^21^ and compares them with those in the HCP842 tractography atlas nonlinearly registered to the subject space^20, 21^. Each track was then labeled by their closest track in the atlas, and the tracks labeled by right or left “vertical occipital fasciculus” were selected (Figure 1). The FA, MD, AD, and RD of bilateral VOF tracts were calculated using built-in functions of the DSI studio.

**Figure 1.**
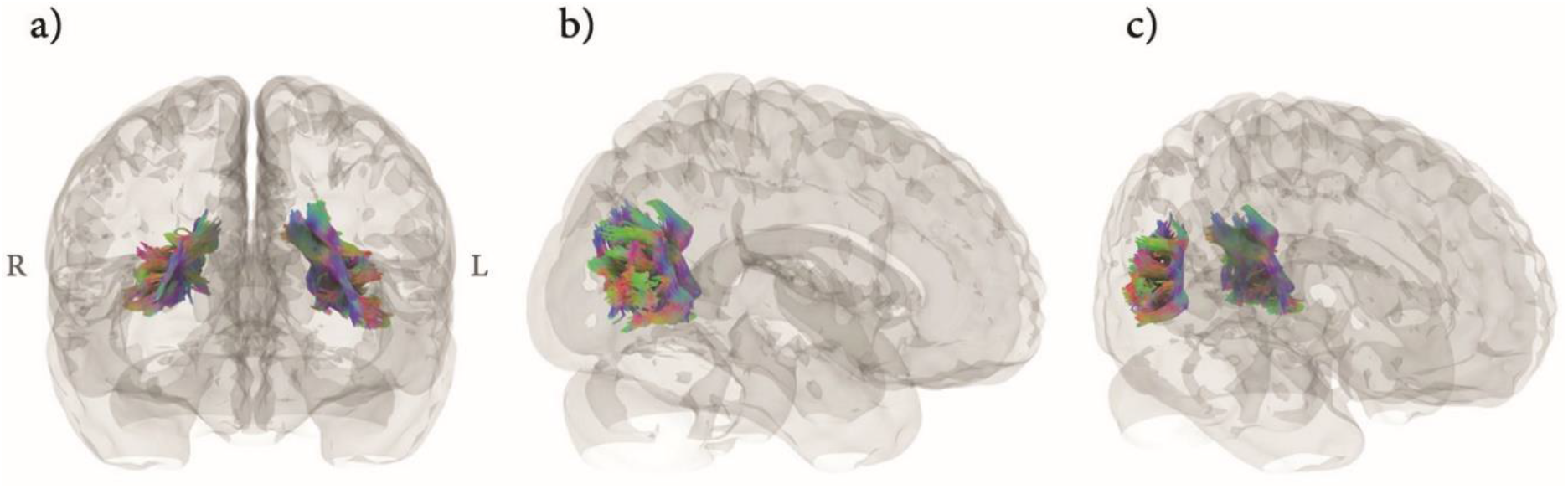
Bilateral Vertical Occipital Fasciculus (VOF) in one of the subjects (ID = MS8003) drawn using automated atlas-based tractography in a) coronal (anterior view), b) sagittal views (right side). In c) the sagittal view is slightly tilted for better visualization of the left VOF. The tracts are directionally color-coded based on the plane they are perpendicular to: red for tracts perpendicular to the sagittal plane, green for tracts perpendicular to the coronal plane, and blue for tracts perpendicular to the axial plane. R = Right, L = Left.

### 2.5. Statistical Analysis

Statistical analysis was conducted using R statistical package^22^ embedded within Rstudio^23^. Students’ t-test or Mann-Whitney U test was done based on the demographics and fiber metrics data distribution to identify between-group differences. We used a general linear model (GLM) with clinical evaluations (EDSS and MSFC) as the outcome variable, the fiber metrics as the predictor, and gender and age as covariates. A normally distributed data of age, BVMT-T, and BVMT-DR was randomly generated in R using the exact number of observations (90 subjects), mean, and standard deviation of these assessments for HC subjects reported in Eshaghi et al.^17^ to compare them with our RRMS group. We also applied GLM on the BVMT-related and EDSS-Vis tests as the outcome variables, and fiber metrics as the predictors to identify the association of fiber integrity with visual disturbances. Another set of the prior GLMs was also used, in which the EDSS was also considered as a covariate to correct for the possible contribution of clinical disease severity on visuospatial learning. Benjamini-Hochberg’s method^24^ was used to correct *p*-values for multiple comparisons wherever applicable.

## 3. Results

### 3.1. Demographics

Age did not follow a normal distribution within groups (Shapiro-Wilk test *p* < 0.001), thus the Mann-Whitney U test was applied. There was no significant difference in age (mean ± SD: RRMS: 31.52 ± 7.82, HC: 30.65 ± 7.82; *p* = 0.52), or gender between groups (Males/Females (M%): RRMS: 12/45 (21.05%), HC: 4/22 (15.38%); *p* = 0.75). The median (and IQR) for disease duration in RRMS group was 6.67 (5.25).

### 3.2. Group Differences in Fiber Metrics

On the left side, RRMS subjects had lower FA but higher MD, AD, and RD compared to HC. On the right side, RRMS subjects had higher MD, AD, and RD compared to HC. There were no significant differences between the number of tracts on either side and FA values of the right VOF. The descriptive statistics and *p*-values are shown in table 2 and figure 2.

**Table 2.**
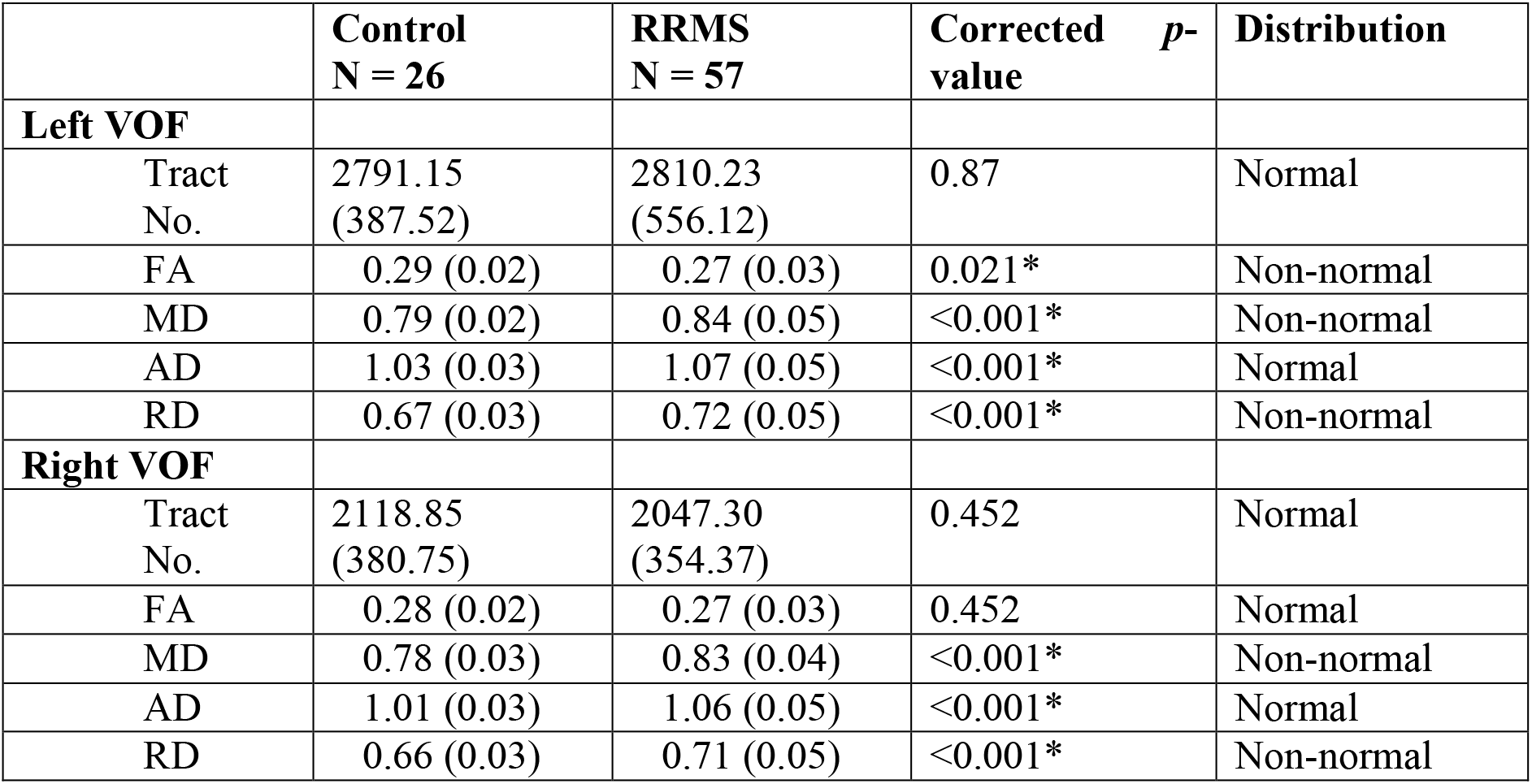
Values of fiber metrics reported as mean ± SD, Benjamini-Hochberg corrected *p*-values, and the data distribution. Distribution was checked using Shapiro-Wilk statistics for fiber metrics in each group, and the distribution was considered non-normal if Shapiro-Wilk *p*-value was less than 0.05 for a fiber metric in each group. Asterisks mark the significant results. VOF = Vertical Occipital Fasciculus, FA = Fractional Anisotropy, MD = Mean Diffusivity, AD = Axial Diffusivity, RD = Radial Diffusivity, Tract No. = Number of Tracts. Significant differences are marked via asterisks.

**Figure 2.**
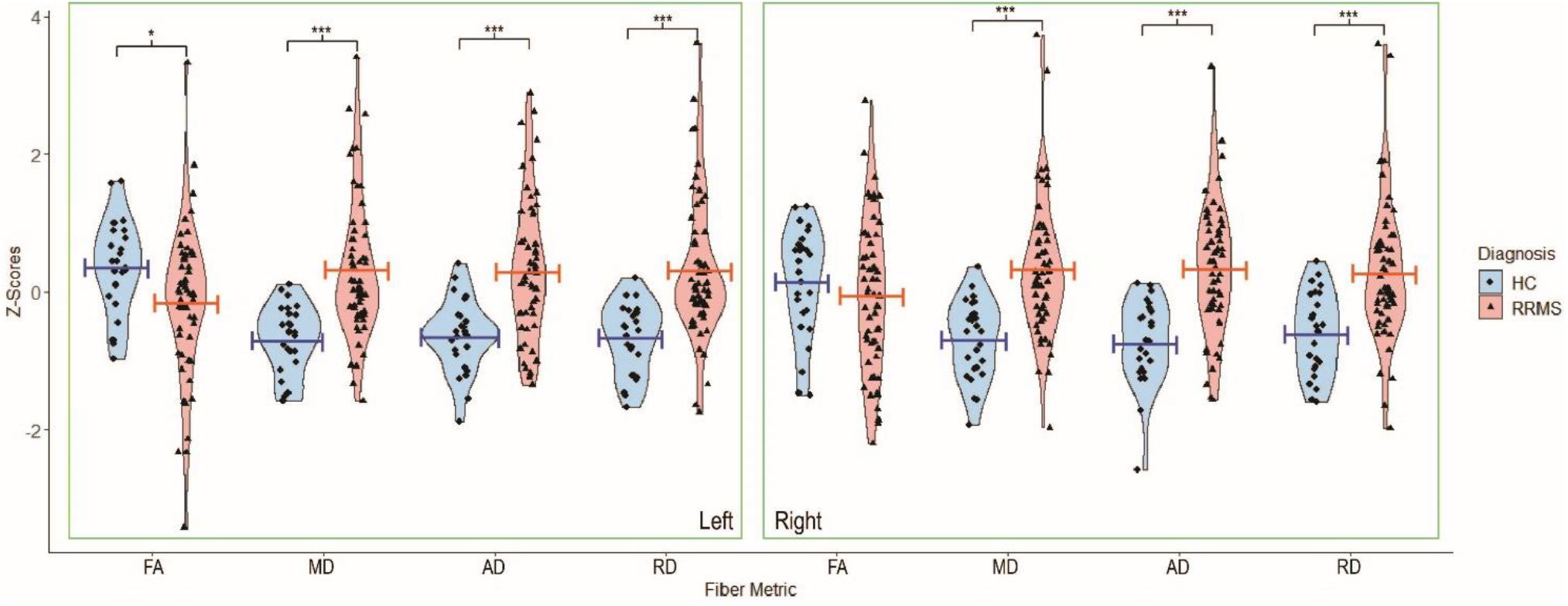
Violin plots showing distribution, mean (lines), and significant differences of fiber metrics based on side and diagnosis group. All values were converted to z-scores for better visualization within one graph. *: *p* < 0.05, ***: *p* < 0.001. HC = Healthy Controls, RRMS = Relapsing-Remitting Multiple Sclerosis, FA = Fractional Anisotropy, MD = Mean Diffusivity, AD = Axial Diffusivity, RD = Radial Diffusivity.

### 3.3. Fiber Integrity and Clinical Evaluation

EDSS data was available for 56 patients (median = 2.5, IQR = 1.5). 53 patients had their PASAT, 9HP, and 25FW tests done, thus MSFC was calculated only for this population (mean ± SD = 0.94 ± 0.16). Both right and left VOF FA were significantly associated with MSFC scores (Estimate (Std. Error): Left FA: -2.39 (0.89), corrected *p* = 0.042; Right FA: - 2.25 (0.84), corrected *p* = 0.042). There were no other significant associations between fiber metrics and clinical evaluations.

### 3.4. Fiber Integrity and Vision

Scores of the visual tests for RRMS patients are available in table 3. There were significant associations between EDSS-Vis and right RD (Estimate (Std. Error) = 8.02 (2.07), corrected *p* = 0.002), right FA (Estimate (Std. Error) = -13.11 (3.71), corrected *p* = 0.003), right MD (Estimate (Std. Error) = 7.09 (2.30), corrected *p* = 0.008), left RD (Estimate (Std. Error) = 5.47 (2.06), corrected *p* = 0.020), and left MD (Estimate (Std. Error) = 5.37 (2.22), corrected *p* = 0.030). There were no significant associations between EDSS-Vis and left FA (Estimate (Std. Error) = -7.44 (3.83), corrected *p* = 0.076), left AD (Estimate (Std. Error) = 3.11 (2.11), corrected *p* = 0.168), and right AD (Estimate (Std. Error) = 2.60 (2.29), corrected *p* = 0.260).

**Table 3.** Values of visual scale of the Expanded Disability Scale Status (EDSS-Vis) and Brief Visuospatial Memory Test – Revised (BVMT-R) for our Relapsing-Remitting Multiple Sclerosis patients. Of note, 56 patients had EDSS and 49 patients had BVMT-R scores. BVMT-DR = BVMT – delayed recall, BVMT-T = BVMT total immediate recall, BVMT-L = BVMT – learning, BVMT-R% = BVMT retained percentage, BVMT-Dis = BVMT discriminability, BVMT-B = BVMT response bias.

| Test | Value |
| --- | --- |
| EDSS-Vis (Count (%)) |  |
| 0 | 33 (59%) |
| 1 | 17 (30.3%) |
| 2 | 4 (7.2%) |
| 3 | 2 (3.5%) |
| BVMT-R (mean (SD)) |  |
| First Trial | 5.53 (2.57) |
| Second Trial | 8.33 (2.87) |
| Third Trial | 9.53 (2.65) |
| BVMT-DR | 9.12 (2.83) |
| BVMT-T | 23.39 (7.53) |
| BVMT-L | 4.12 (1.74) |
| BVMT-R% | 93.28 (13.62) |
| BVMT-Dis | 4.75 (2.16) |
| BVMT-B | 0.45 (0.13) |

49 patients had their BVMT evaluations completed. Before comparing the difference between our RRMS sample and HC from Eshaghi et al. (“HC” from now on only in the current paragraph), we evaluated the difference between the demographics of the two samples. They have recruited 90 HC subjects (33 males, 57 females) with the mean age of 33.65 (±9.48)^17^. Gender distribution between HC and RRMS differ significantly (Males/Females (M%): HC = 33/57 (36.67%), RRMS = 12/45 (21.05%), *p* = 0.045). Since age, BVMT-T, and BVMT-DR did not follow normal distribution in our sample (all Shapiro-Wilk *p* < 0.05), we used Mann-Whitney U test for the related comparisons. Age was significantly different between the two groups (HC vs. RRMS (mean (SD)): 33.65 (9.48) vs. 31.52 (7.82), *p* = 0.037). Comparing the BVMT-T and BVMT-DR between the two groups, no significant differences were noted (BVMT-T: HC vs. RRMS (mean (SD)): 25.01 (7.38) vs. 23.39 (7.53), *p* = 0.70; BVMT-DR: HC vs. RRMS (mean (SD)): 9.59 (2.76) vs. 9.12 (2.83), *p* = 0.55).

After adjusting for covariates and multiple comparisons, there were significant associations between BVMT-T and left MD (Estimate (Std. Error) = -78.68 (20.17), corrected *p* = 0.002), left RD (Estimate (Std. Error) = -73.49 (19.65), corrected *p* = 0.002), left AD (Estimate (Std. Error) = -66.68 (19.14), corrected *p* = 0.002), and right AD (Estimate (Std. Error) = -57.39 (21.94), corrected *p* = 0.024). There were also significant associations between BVMT-DR and left MD (Estimate (Std. Error) = -26.13 (8.05), corrected *p* = 0.011), left AD (Estimate (Std. Error) = -22.67 (7.54), corrected *p* = 0.011), and left RD (Estimate (Std. Error) = -24.12 (7.84), corrected *p* = 0.011). After adding EDSS as a covariate, the same associations remained significant with the same directionality of the estimate (Negative associations of BVMT-T with left MD, RD, AD, and right AD, and negative associations of BVMT-DR with left MD, AD, and RD; all corrected *p*s < 0.042), with the addition of two new significant associations: BVMT-Dis and right MD (Estimate (Std. Error) = 17.05 (6.63), corrected *p* = 0.052) and right RD (Estimate (Std. Error) = 16.37 (6.21), corrected *p* = 0.052). No other significant associations were noted.

## 4. Discussion

This is the first study investigating microstructural changes of VOF in MS patients to the best of our knowledge. Our results showed higher MD, AD, and RD in bilateral VOFs and lower FA in the left VOF in our RRMS population compared to healthy controls, implying lower integrity of VOF in RRMS. Our results also showed a negative association between FA and MSFC in bilateral VOFs. To investigate the association between visual performance and microstructural integrity of white matter in VOF, we showed poorer function in visual part of EDSS, lower total immediate recall, and lower delayed recall of BVMT were associated with lower integrity of right and left VOFs.

Recent discoveries show that VOF is located in the posterolateral region of the brain, lying lateral to inferior longitudinal and inferior frontal occipital fasciculus, and posterior to arcuate fasciculus^8^. Besides cortical projections of VOF into the lateral occipital cortex (e.g., LO-1 and LO-2)^25^, this pathway projects into the ventral and dorsal visual cortex^26^. Ventral projections of VOF end in the inferior occipital gyrus, inferior occipital sulcus, and posterior transverse collateral sulcus, where VO-1/hV4 visual areas are located^26^. Dorsal projections of VOF terminate in the posterior intraparietal sulcus and transverse occipital sulcus, where V3A/B visual areas are located^8^.

Disrupted myelination is observable within the white matter in MS due to prior lesions or an on-going heightened neuroinflammatory responses^1^. MS was one of the pioneer disorders in deploying DWI to study white matter integrity due to its underlying pathological mechanisms^27^. Prior studies have shown disrupted integrity in the tracts traversing the white matter, such as optic radiation and optic nerve, and their association with clinical status^2, 3^. Thus, it was not surprising that we found decreased fiber integrity in bilateral VOFs in our sample compared to the healthy controls. We also showed worse clinical outcomes based on MSFC are associated with VOF integrity, reflecting general neuroinflammation within the brain causing damage to white matter, including VOF.

Based on its anatomical connections, it is expected that damages to VOF can cause disturbances in visual functions. On the other hand, fMRI studies suggest that interpretation of particular visual information sets necessitates concurrent activation of ventral and dorsal visual streams^28^. Although the disturbances in anterior visual pathways and optic radiation in MS are widely investigated in the literature and might account for the visual disturbances in MS patients^29^, we propose for the very first time that the microstructural disintegration of VOF in MS patients interrupts the communication between different areas of the vision-associated cortex and leads to poor performance in visual tests including BVMT and EDSS-Vis. Interestingly, this finding was independent of the disease severity based on EDSS total score.

The reported selective association between fiber integrity and total immediate and delayed recall parts of BVMT (BVMT-T and BVMT-DR), and not other measures (such as learning, retained percentage, discriminability, and bias), is an interesting finding. Proper performance in BVMT requires the appropriate function of both visuospatial and learning cognitive functions, either separately and coordinated^16^. These two functions roughly relate to dorsal and ventral visual streams, respectively. The dorsal stream connects to parietal and frontal cortices, answering the question of “Where?”, coordinating attention and vision-related motions, wherever the ventral stream tries to answer the “What?” question by traversing through the temporal lobe^10, 11, 30^. In this regard, it seems there are two separate visual working memory systems: the “object” working memory, which is more dependent on “What?” areas in the temporal cortices (e.g., superior temporal gyrus) and “spatial” working memory which is more dependent to “Where?” stream and prefrontal working memory (e.g., dorsolateral prefrontal cortex)^31, 32^. Also, both the drawn object and its location (i.e., spatial) are essential for scoring BVMT. The natures of the recall parts of BVMT are visual working memory task, requiring the integration between object-based and spatial visual working memory systems to function correctly. Thus, we propose that since BVMT-T and BVMT-DR are highly dependent on proper communication of ventral and dorsal streams, they are strongly related to the VOF integrity.

We did not have BVMT scores for HCs om the CRIMSON dataset, so we compared BVMT-T and BVMT-DR of our RRMS sample with HCs from another dataset acquired at the same research center two years earlier^17^. Despite our expectations, there were no significant differences in BVMT-T and BVMT-DR between our RRMS and their HC subjects. No differences in these measures (esp. BVMT-T) between RRMS and HC have also been reported in other studies^33, 34^. Interestingly, HCs had higher BVMT-T and BVMT-DR than MS subjects in Eshaghi et al. This incongruence in the results can be attributed to a few factors. First, they have included RRMS and secondary progressive MS subjects in their MS group. Moreover, their HC subjects were older than our RRMS patients, potentially affecting the results due to the detrimental effects of normal aging on BVMT measures^16^.

Despite using a sophisticated imaging analysis pipeline and a sensitive task to evaluate visuospatial learning and memory, our study is faced with limitations. Due to the hospital-setting and data collection of the CRIMSON study, DWI scans were done with a 1.5T MRI scanner with one b-value. It has been shown multi-shell DWI acquisitions can better identify fibers crossing over each other^4, 35^. The position of VOF and proximity to inferior longitudinal and inferior frontal occipital fasciculus necessitates addressing fiber cross-over issue^8^. We used high number of diffusion gradient directions (n = 64) and relatively high b-value (b = 1000 s/mm^2^) that can somehow address this issue. However, studies using a higher sample-size, implementing visuospatial learning tasks to patients with variable disease course (addition of secondary or primary progressive MS subjects) and healthy controls, and acquiring DWI with multi-shell protocols may further clarify the picture provided in this article.

In conclusion, this study provides evidence for decreased microstructural integrity of VOF in MS patients. Moreover, DWI metrics of white matter integrity were associated with clinical status, visual function, and visuospatial learning, independent of covariates such as disease severity. Regarding the role of VOF in functional and structural connectivity of ventral and dorsal visual streams, the disrupted integrity of VOF probably underlies the visual dysfunction in MS patients.

## Data Availability

The imaging analysis pipeline of this project is based on the cloud computing platform brainlife.io which can be accessed upon request. Moreover, the codes used to run the analysis on R are available upon request from the corresponding author. The data used in this project were from the CRIMSON study. To access that an email must be sent to the lead investigators of that study, available in the manuscript [ref. 13].

## Acknowledgments

We thank Dr. Franco Pestilli, developers and contributors of the Brainlife.io platform, and Dr. Reza Rajimehr for his help due to his extensive knowledge and expertise in vision science.

